# Demand for Self-Managed Online Telemedicine Abortion in Eight European Countries During the COVID-19 Pandemic: A Regression Discontinuity Analysis

**DOI:** 10.1101/2020.09.15.20195222

**Authors:** Abigail R. A. Aiken, Jennifer E. Starling, Rebecca Gomperts, James G. Scott, Catherine E. Aiken

**Affiliations:** Associate Professor, LBJ School of Public Policy, University of Texas at Austin, Texas, 78712, USA; Faculty Associate, Population Research Center, University of Texas at Austin, Texas, 78712, USA; PhD Candidate, Department of Statistics and Data Sciences, University of Texas at Austin, Austin, TX 78712, USA; Statistician, Mathematica Policy Research, Cambridge, MA, 02139, USA; Founder and Director, Women on Web, Amsterdam, The Netherlands; Professor, McCombs School of Business, University of Texas at Austin, Austin, TX, 78712, USA; Academic Clinical Lecturer, University Department of Obstetrics and Gynaecology, University of Cambridge; NIHR Cambridge Biomedical Research Centre, CB2 2SW, UK

## Abstract

**Objectives:** In most European countries, patients seeking medication abortion during the COVID-19 pandemic are still expected to attend healthcare settings in person despite lockdown measures and infection risk. We assessed whether demand for self-managed medication abortion provided by a fully remote online telemedicine service increased following the emergence of COVID-19.

**Design:** We used regression discontinuity to compare the number of requests to online telemedicine service Women on Web in eight European countries before and after they implemented lockdown measures to slow COVID-19 transmission. We examined the number deaths due to COVID-19, the degree of government-provided economic support, the severity of lockdown travel restrictions, and the medication abortion service provision model in countries with and without significant changes in requests.

**Setting:** Eight European countries served by Women on Web.

**Participants:** 3,915 people who made requests for self-managed abortion to Women on Web between January 1^st^, 2019 and June 1^st^, 2020.

**Main Outcome Measures:** Percent change in requests to Women on Web before and after the emergence of COVID-19 and associated lockdown measures.

**Results:** Five countries showed significant increases in requests, ranging from 28% in Northern Ireland (p=0.001) to 139% in Portugal (p<0.001). Two countries showed no significant change in requests, and one country, Great Britain, showed an 88% decrease in requests (p<0.001). Countries with significant increases in requests were either countries where abortion services are mainly provided in hospitals or where no abortion services are available and international travel was prohibited during lockdown. By contrast, Great Britain authorized teleconsultation for medication abortion and provision of medications by mail during the pandemic.

**Conclusion:** These marked changes in requests for self-managed medication abortion during COVID-19 demonstrate demand for fully remote models of abortion care and an urgent need for policymakers to expand access to medication abortion by telemedicine.

What this paper adds

What is already know on this subject

- The COVID-19 pandemic has presented challenges to patients seeking medication abortion, including lockdown travel restrictions and infection risk during in-person clinic visits.
- Yet in most European countries, medication abortion must still be provided through in-person models of care. The sole exception is Great Britain, where a fully remote medication abortion service was introduced in response to the pandemic.
- Anecdotal reports suggest that patients are struggling to access in-person abortion services and may turn to self-managed abortion as a result. However, to date there has been no systematic assessment of this possibility.

What this study adds

- Our study provides the best evidence to date that demand for self-managed medication abortion provided using online telemedicine increased following the emergence of the COVID-19 pandemic.
- The largest increases were observed in countries where medication abortion is provided mainly in hospitals and where travel restrictions were most stringent. By contrast, in the one country that implemented fully remote services, demand for self-managed abortion declined almost to zero.
- Our findings demonstrate the urgent need for policymakers to expand access to telemedicine models of medication abortion within the formal healthcare setting.

## Introduction

The first wave of the COVID-19 pandemic posed challenges for the provision of abortion care in Europe. Reallocation of resources, redeployment of staff, and social distancing requirements all introduced new barriers to in-person clinic visits.^1,2^

Countries differed in their policy responses to these new challenges. Great Britain expanded remote access to medication abortion, allowing teleconsultation with providers, and mifepristone and misoprostol to be provided by mail.^3-5^ France extended the ability to take abortion medications at home following an in-person visit with a healthcare professional from 7 weeks to 9 weeks of gestation.^6^ Germany allowed mandatory pre-abortion counselling to take place by phone or video teleconsult instead of in person.^7^ Most other countries, however, made few changes to medication abortion service models and continued to require fully in-person provision, despite calls from human rights groups to prioritize patient safety and expand remote access.^7,8^

At the same time, the economic downtown and rising unemployment across Europe in the wake of the pandemic may increase demand for abortion care at a time when it is most difficult to access in the clinic setting. This situation raises the possibility of an increase in abortions taking place outside the formal healthcare setting. Marie Stopes International estimates that an additional 2.7 million unsafe abortions will take place globally as a result of health service disruptions caused by the pandemic.^9^

However, not all abortions that take place outside the formal healthcare settings are unsafe. Medication abortion provided through online telemedicine services has been shown to be a safe and effective option.^10,11^ Using data from one such service, we assessed whether demand for online telemedicine abortion changed in eight European countries after stay-at-home restrictions to slow the spread of COVID-19 were introduced.

## Methods

We obtained fully de-identified data from Women on Web (WoW), a non-profit organisation that provides telemedicine medication abortion services up to 10 weeks of gestation.^12^ The service is accessed via an online form, which directly populates the database from which our data were obtained. Submitted forms are screened by a doctor, and if clinical eligibility criteria are met, mifepristone and misoprostol are sent by mail. In some countries, referrals are also made to local in-clinic services. A donation of 70-90 Euros is requested to support the service, but may be waived or reduced in cases of financial hardship. Information and support are provided via email in a variety of languages by a trained helpdesk team. People accessing the service consent to the fully anonymized use of their data for research purposes at the time of submitting the online consultation form.

Our analytic sample includes eight countries: Germany, Hungary, Italy, Malta, The Netherlands, Northern Ireland, Portugal, and Great Britain. WoW does not accept consultations from all countries in Europe, because abortion is legal and normally relatively accessible in most places. Among those countries that WoW does serve, some have only a few consultations requests over the course of a year. We excluded countries that had too few requests to reliable detect differences in request numbers between the ‘before’ and ‘after’ periods (i.e. fewer than 10 expected requests in the ‘after’ period). We also excluded Spain, because the Spanish Government censored the WoW website during the study period and so no requests could be made,^13^ and Poland because the number of requests made to WoW has been unstable since the beginning of 2020.

We obtained the daily number of requests made to WoW from the eight countries in our sample between 1^st^ January 2019 and 1^st^ June 2020 (the last day that lockdown measures were lifted in a country included in the analysis). We excluded duplicate requests, which were identified as >1 request with the same information and location made within 12 hours. The number of requests from each country was analysed using a regression-discontinuity design.^14^ We designated a ‘before’ period, which began on 1^st^ January 2019 and ended on the date that each individual country’s government issued their first ‘stay-at-home’ directive. The one exception was Germany, where the ‘before’ period begins on 1^st^ January 2020, due to the fact that WoW did not accept consultations from Germany in until late 2019. The ‘after’ period began the first day after the ‘stay-at-home’ directive was issued for each country, and ended on the first day that the directives were eased in each country. ‘Stay-at-home’ directives were chosen as the threshold date defining the “pre” and “post” periods, because the majority of European countries issued such a directive, which posed definitive limitations on population movement and activities. Of the countries included in our analytic sample, only Malta did not issue a population-wide directive, and we instead used the date that the Maltese government issued a directive to close public places as the discontinuity point.^15^

We fit a generalised linear model (GLM) for each country’s daily requests between 1^st^ January 2019 (1^st^ January 2020, for Germany) and the date of easing ‘stay-at-home’ restrictions. The model incorporated a dummy variable for the ‘before’ v. ‘after’ period, representing a possible discontinuity at the day of the ‘stay-at-home’ directive. The significance of the discontinuity for each country was assessed using a likelihood ratio test to compare with a null model that did not include a dummy variable for the ‘before’ v. ‘after’ period. The null model was also used to generate Monte Carlo simulations for each country, which create a probability distribution of the expected requests in the ‘after’ period with no discontinuity. The observed requests line would be highly likely to lie within this probability distribution if there was no difference in requests between the ‘before’ and ‘after’ periods. We also calculated the percentage difference between observed and expected requests in the ‘after’ period. For Northern Ireland, both the null and discontinuity models also included a dummy variable indicating the period after 10^th^ April 2020, because abortion services became available for the first time in Northern Ireland on this date.^16^

We also compiled information for each country included in the analysis on several metrics we hypothesised could be related to demand for online abortion: stringency of ‘stay-at-home’ requirements; deaths due to COVID-19; economic assistance provided by governments in response to the pandemic; ^17^ and abortion service provision before and during the pandemic.^7,18-20^ We examined each of these metrics across each country included in the analysis to assess their relationship to changes in requests to WoW.

Data analyses were conducted using the R statistical package version 3.6.2.^21^ Findings were considered statistically significant at an alpha level of 0.05. The study was reviewed by the University of Texas at Austin Institutional Review Board and considered exempt on the basis that the study is an analysis of pre-collected, fully de-identified data.

### Patient involvement

Patients were not involved in the design or conduct of the study. However, the follow-up that WoW provides is designed to address the priorities and experiences of people who access the service. Thus, although this study is an analysis of secondary, de-identified data, with no direct participant involvement, the research questions were informed by the needs of people who rely on WoW to access abortion.

## Results

During the data collection period, Women on Web received 3,915 requests for abortion medications from the eight countries included in the analysis. Among these, we observed a statistically significant increase in requests during the ‘after’ period in five countries: Hungary, Italy, Malta, Portugal, and Northern Ireland (Figure 1). The magnitude of the observed increases ranged from 139% above expected in Portugal to 28% above expected in Northern Ireland (Table 1). In two countries (Germany, the Netherlands) there was no statistically significant difference in observed compared to expected numbers of requests in the ‘after’ period (Figure 1 and Table 1). In one country (Great Britain), there was a statistically significant decrease in requests in the ‘after’ period (Figure 1 and Table 1).

**Table 1:** Actual versus expected numbers of self-managed abortion requests in the “after” period for each country included in the study.

| Country | Actual Requests | Expected Requests | Percent Change Over Baseline Trend (95% CI) | P Value |
| --- | --- | --- | --- | --- |
| Portugal | 34 | 14.2 | 139.0<br>(54.5, 385.7) | < 0.001 |
| Italy | 53 | 31.6 | 67.9<br>(23.3, 152.4) | < 0.001 |
| Hungary | 113 | 83.2 | 35.8<br>(11.9, 71.2) | < 0.001 |
| Malta | 69 | 52.3 | 31.9<br>(3.0, 76.9) | < 0.001 |
| Northern Ireland (UK) | 97 | 75.8 | 28.0<br>(4.3, 64.4) | 0.001 |
| Germany | 465 | 467.1 | -0.5<br>(-9.0, 9.2) | 0.798 |
| Netherlands | 47 | 50.9 | -7.7<br>(-28.8, 27.0) | 0.458 |
| Great Britain | 1 | 8.1 | -87.6<br>(-92.9, -66.7) | < 0.001 |

**Figure 1:**
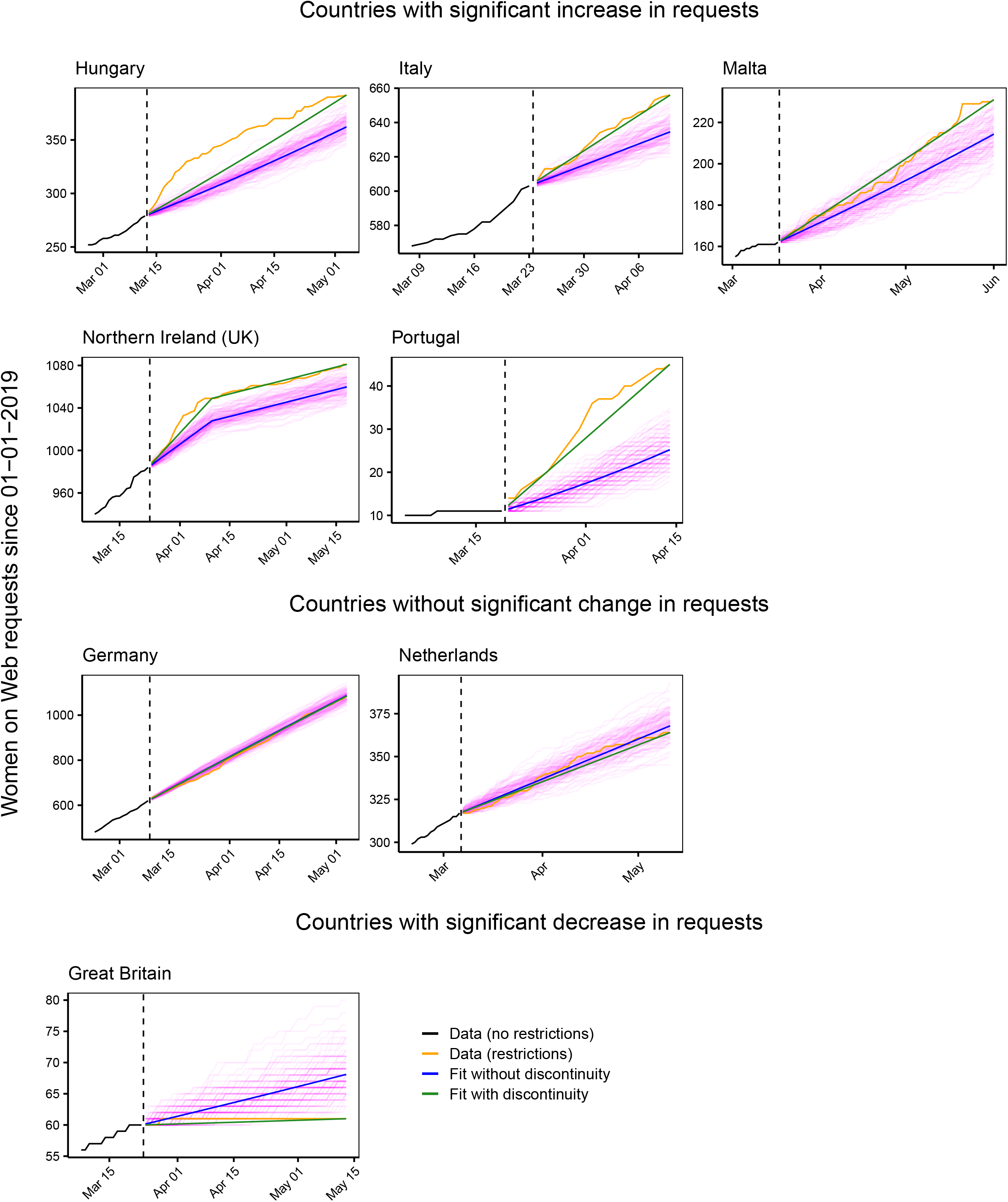
Observed versus expected requests to Women on Web for all countries included in the analysis. Cumulative requests in the “before” versus “after” periods are in black and orange, respectively. Vertical dashed lines show the dates when stay-at-home orders were announced. The blue line shows the model without any discontinuities (the null model), and the green line shows the model fit with a discontinuity. for the stay-at-home order. The pink lines are the 250 Monte Carlo simulations from the null model, which support the likelihood ratio test’s finding that the model with discontinuities is a significantly better fit than the null model.

Countries that had higher numbers of COVID-19 related deaths or which provided less government economic support during the pandemic did not appear to have higher numbers of requests to WoW (Table 2). We did however, observe a relationship between higher numbers of requests and both the location of abortion service provision, and the severity of domestic and international travel restrictions (Table 2). In Italy, Portugal, and Hungary, all of which showed significant increases in requests to WoW, abortion is provided mostly in the hospital setting and all enacted stringent stay-at-home requirements. In Northern Ireland and Malta, where significant increases in requests were also observed, in-clinic abortion services are only available by traveling outside of the country, and international travel was restricted during the study periods. In Germany and the Netherlands, we observed no increases in requests, abortion services remained available in clinic settings, and no country-wide domestic travel restrictions were enacted. In Great Britain, abortion services were made available by fully remote telemedicine shortly after lockdown began and we observed a significant decrease in requests.

**Table 2:** Financial and health parameters during the COVID-19 pandemic for all countries included in the analysis.

| Country | Date stay-at-home requirements began | Date stay-at-home requirements ended | Number of Deaths due to COVID-19 when stay-at-home requirement ended | Stringency of stay-at-home requirements (Country-wide domestic travel restriction) | Government economic support index | Location and scope of abortion service provision |
| --- | --- | --- | --- | --- | --- | --- |
| Portugal | 19/03/20 | 14/04/20 | 535 | 87.96<br>(Yes) | 75 | Abortion available on request through 10 weeks gestation with a 3-day waiting period and provided mostly in hospitals. |
| Northern Ireland | 23/03/20 | 10/04/20 | 476 | 75.93<br>(Yes, including to the rest of the UK) | 100 | No abortion services available until new legislation brought into effect on April 10<sup>th</sup> 2020. |
| Hungary | 12/03/20 | 04/05/20 | 351 | 76.85<br>(Yes) | 75 | Abortion available in certain circumstances through 12 weeks gestation and provided in hospitals. No medication abortion available. |
| Malta | 17/03/20<sup>†</sup> | 01/06/20 | 9 | NA<br>(No, but no travel outside the country) | NA | No abortion services available. |
| Italy | 23/03/20 | 10/04/20 | 18,281 | 93.52<br>(Yes) | 50 | Abortion available on request through 90 days gestation with a 7-day waiting period, and provided mostly in hospitals. |
| Germany | 09/03/20 | 04/05/20 | 6,692 | 73.15<br>(No, only a few specific districts) | 88 | Abortions provided on request through 14 weeks with a 3-day waiting period, and mostly provided in doctor's offices and clinics. |
| Netherlands | 06/03/20 | 11/05/20 | 5,440 | 80<br>(No) | 63 | Abortions provided on request up until viability with a 5-day waiting period, and mostly provided in doctor's offices and clinics. |
| Great Britain | 23/03/20 | 13/05/20 | 32,692 | 76<br>(Yes) | 100 | Medication abortion provision changed from in—person to fully remote service model, including phone consultation and pills provided by mail or pick-up at a clinic |

<sup>†</sup> Malta did not issue a population-wide directive, so that date that the Maltese government issued a directive to close public places is used in lieu.

## Discussion

During the first wave of the COVID-19 pandemic in Europe, we observed changes in requests to the WoW online telemedicine abortion service among five out of eight countries in our analysis. Among countries where abortion is legally available within the formal healthcare setting, we observed increased requests to the WoW online telemedicine service in those countries that had more stringent stay-at-home requirements––including country-wide domestic travel restrictions––and where abortion is mostly available only in the hospital setting. Among the two countries where abortion was not legally available within the formal healthcare setting during the study period, and where travel outside the country was restricted, we also observed an increase in requests. Among countries where abortion is legally available but which enacted less stringent stay-at-home policies (including no country-wide domestic travel restrictions) and where abortions are provided outside the hospital setting, we observed no increases in requests. In the sole country where abortion services were made available by fully remote telemedicine during the study period, we observed a significant decrease in requests.

Our data provide a unique window into requests for self-managed medication abortion using online telemedicine during COVID-19. Key strengths include the ability to measure changes in demand for self-managed abortion from a reliable source that does not rely on self-reporting, and the ability to compare data from before and after the emergence of COVID-19. An important limitation, however, is that there are of course other pathways to abortion outside the formal healthcare setting in Europe, including alternative sources of mifepristone and misoprostol, and non-medication methods. Thus, we cannot measure all demand for self-managed abortion during the pandemic. We also lack nuanced insight into the exact reasons underlying changes in requests to WoW for any particular country. It is important to note that in the two countries where we observed no increases in requests, people likely still encountered challenges to accessing abortion services.^22^ Future qualitative work could address this important knowledge gap.

Our results may reflect two distinct phenomena. First, in some countries, more people may be seeking abortion through all channels during the pandemic. The decision to end a pregnancy could be due to the perception of risk posed by COVID-19, reduced access to pre-natal care, and limited social support during lockdowns.^23^ Additionally, decision-making could be influenced by the economic downturn COVID-19 has precipitated, with many people facing unemployment or financial losses.^24^ It has also been suggested that social distancing policies may increase rates of unintended pregnancy due to increased time spent at home with a partner or reduced access to contraception.^25^

Second, the observed increases in requests may represent a shift in demand from in-clinic abortion to self-managed abortion using online telemedicine. In countries where abortion services are provided predominantly or solely in hospital settings, people may have feared entering a hospital due to perceived or real risk of infection. Even where limited alternatives are available in the community setting––for example in Portugal, where a few private clinics offer abortion services––accessing these services may still have been extremely challenging due to the infection risk associated with public transport, inability to escape surveillance from a controlling partner, or difficulty finding childcare while daycares and schools were closed. Moreover, in countries with no abortion services, the inability to travel outside of the country to seek abortion care due to travel restrictions may have led more people to seek an alternative in online telemedicine. Indeed, our findings from Northern Ireland show a steep increase in requests to WoW following the introduction of lockdown measures, followed by a levelling off shortly after the introduction of within-country abortion services.

These challenges to accessing medication abortion during COVID-19, coupled with the increases we observed in requests for online telemedicine are reflected in the fact that people in countries where the challenges are greatest found their own solutions outside the clinic setting. However, while medication abortion provided via online telemedicine is a safe and effective option,^10,11^ it is not without legal risks.^26^ Its safety also depends on access to the formal healthcare system when necessary, which is not guaranteed during a pandemic. Additionally, while some people may prefer self-managed medication abortion, others may experience it as fraught and isolating due to stigma or the threat of criminalization, or may have preferred in-clinic care.^27^ Despite the fact that the WHO recommends the use of telemedicine abortion provision models during COVID-19,^28^ only one country in the analysis, Great Britain, responded to pandemic by purposefully changing their medication abortion service to circumvent the difficulties of in-person care. Following the introduction of a fully remote telemedicine service for medication abortion up to 10 week’s gestation, where consultations with healthcare professionals are done by phone or video, and medications are mailed or made available for pick-up from a clinic front-desk, requests to WoW decreased to a single consultation. This dramatic decrease points not only to the removal of access barriers posed by COVID-19, but also of pre-existing barriers. Evidence from other settings suggests that similar telemedicine models for medication abortion are safe, effective, and acceptable to patients.^29^

## Conclusion

Our findings provide evidence in support of the need for service model changes to make medication abortion more accessible during and beyond the COVID-19 pandemic.^30^ Fully remote provision of early medication abortion negates the need to visit a hospital or healthcare facility, thus preserving personal protective equipment, and reducing infection risks for both patients and healthcare providers. Follow-up care can be provided in the clinic if necessary, and patients have clear continuity of care in the rare instances that adverse events occur. Authorizing and implementing telemedicine models within the formal healthcare setting in line with the WHO recommendations would help to meet the demand we observed for remote provision and would ensure truly patient-centered care.

## Data Availability

No additional data are available.
All authors, external and internal, had full access to all of the data (including statistical reports and tables) in the study and can take responsibility for the integrity of the data and the accuracy of the data analysis.

## Contributors

ARAA and CEA conceived of the original research question. ARAA, CEA, JGS, and JES contributed to the study design. RG provided the de-identified data. JES conducted the statistical analyses and prepared the tables and figures. ARAA and CEA did the initial data interpretation. ARAA wrote the first draft of the manuscript. All authors contributed to final data interpretation, revised first and subsequent drafts critically for intellectual content, and approved the final manuscript. All authors agree to be accountable for all aspects of the work. ARAA is the manuscripts guarantor. The corresponding author attests that all listed authors meet authorship criteria and that no others meeting the criteria have been omitted.

## Copyright

The Corresponding Author has the right to grant on behalf of all authors and does grant on behalf of all authors, a worldwide licence to the Publishers and its licensees in perpetuity, in all forms, formats and media (whether known now or created in the future), to i) publish, reproduce, distribute, display and store the Contribution, ii) translate the Contribution into other languages, create adaptations, reprints, include within collections and create summaries, extracts and/or, abstracts of the Contribution, iii) create any other derivative work(s) based on the Contribution, iv) to exploit all subsidiary rights in the Contribution, v) the inclusion of electronic links from the Contribution to third party material where-ever it may be located; and, vi) licence any third party to do any or all of the above.

## Data Sharing

No additional data are available.

All authors, external and internal, had full access to all of the data (including statistical reports and tables) in the study and can take responsibility for the integrity of the data and the accuracy of the data analysis.

## Ethics Approval

The University of Texas at Austin Institutional Review Board reviewed the study protocol and declared the use of pre-collected, fully de-identified data exempt from the need for approval.

## Transparency

ARAA affirms that the manuscript is an honest, accurate, and transparent account of the study being reported; that no important aspects of the study have been omitted; and that any discrepancies from the study as planned (and, if relevant, registered) have been explained

## Dissemination Declaration

We plan to disseminate the results to people who have made requests to the WoW service by having a link to the published paper included in the ‘Research’ section of the WoW website.

